# Intimate partner violence and its correlates in middle-aged and older adults during the COVID-19 pandemic: A multi-country secondary analysis

**DOI:** 10.1101/2023.09.26.23296197

**Authors:** Gwendolyn Chang, Joseph Tucker, Kate Walker, Claire Chu, Naomi Miall, Rayner Tan, Dan Wu

## Abstract

**Background:** Intimate partner violence (IPV) may have been exacerbated during the COVID-19 pandemic. Middle-aged and older adults, ages 45 years or older, are at higher risk of COVID-19 mortality and social isolation. However, most studies on IPV during the pandemic do not focus on this important subpopulation. Informed by the social-ecological theory, this study examines individual, household, community, and country-level correlates of IPV among middle-aged and older adults in multiple countries using a cross-sectional online survey.

**Methods:** Data from 2867 participants aged over 45 in the International Sexual Health and Reproductive Health (I-SHARE) survey from July 2020 to February 2021 were described using univariate analysis. IPV was defined using four validated WHO measures. Individual characteristics included self-isolation and food security. At the country-level, we examined social distancing stringency. Logistic regression models with a random intercept for country were conducted to explore IPV correlates among 1730 eligible individuals from 20 countries with complete data.

**Results:** Most participants were heterosexual (2469/2867), cisgender (2531/2867) females (1589/2867) between the ages of 45-54 (1539/2867). 12.1% (346/2867) of participants experienced IPV during social distancing measures. After adjustment, participants who self-isolated experienced 1.4 (95% CI 1.0, 2.0, p=0.04) times the odds of IPV compared to those who had not isolated. Those who reported an increase in food insecurity compared to pre-pandemic experienced 2.2 times the odds (95% CI 1.6, 3.0, p<0.0001) of IPV compared to those who did not report increased food insecurity. People in countries with more stringent social distancing policies were less likely to experience IPV compared to people in countries with lower levels of stringency (aOR=0.6, 95% CI 0.4, 0.9, p=0.04).

**Conclusions:** IPV was common among middle-aged and older adults during the COVID-19 pandemic. Our data suggest the need for further crisis management and social protection measures for middle-aged and older adults who have intersecting vulnerabilities to IPV to mitigate COVID-19 impact.

## Background

Intimate partner violence (IPV) is defined by the World Health Organization (WHO) as “any behaviour within an intimate relationship that causes physical, psychological, or sexual harm to those in the relationship” by current or former partners(1,2). IPV is a pervasive public health issue with an estimated global lifetime prevalence of 30% among women and 10% among men (3,4). IPV is also a growing problem that intersects with population aging (5). Middle-aged and older people are at higher risk of disability, social isolation, chronic illness, and cognitive impairment, which make them more vulnerable to new or worsening forms of violence (6). But previous literature on IPV primarily focuses on younger people (7), neglecting the unique experiences among middle-aged and older adults, such as increasing dependence on partners as caregivers and menopause (8). A nationally representative survey conducted in the United States of over 5,000 older adults (2009) found nearly 10% of older adults faced some form of violence or abuse and that partners and spouses were the perpetrators of more than half of the reported violence (9). This analysis focused on adults aged 45 or older, capturing middle-aged and older people.

Previously reported correlates of IPV were operationalised at four levels based on an adapted social-ecological theory (2006) (10). These were individual-level sociodemographic and behavioral characteristics; relationship/household level factors capturing characteristics of intimate partners, family members, and peers; community contexts such as residential areas; and larger country-level societal factors including socioeconomic conditions, gender inequity, social policies (10). Violence is a complex interplay between each of these levels. This multi-level theory has been used to understand different types of violence, including IPV (11) and elder abuse (12). Understanding factors at each of these levels is important in public health to identify populations at risk and develop interventions and policies that address violence at different levels (10,11).

Since the COVID-19 pandemic, there have been concerns that widespread social distancing measures could put more people at risk of IPV (5,13,14). The pandemic may have exacerbated some IPV risk factors such as dependency on others for care (15). While social distancing measures effectively reduced COVID-19 infections, these measures may have unintended consequences that increased IPV risk. There is limited multi-country data on IPV risk during COVID-19. No studies have focused on IPV among middle-aged and older adults during COVID-19. More research is needed to understand the unintended consequences of social distancing policies and their potential ongoing and long-lasting consequences on IPV.

This secondary analysis seeks to address the gap in literature by analysing correlates of IPV during COVID-19 in middle-aged and older adults using the International Sexual Health and Reproductive Health in the times of COVID-19 (I-SHARE 2020-21) survey data. The I-SHARE 2020-21 survey is a multi-country online survey that harmonises sexual and reproductive health instruments for global comparison using an open science approach (16). This analysis was used to research multi-country and multi-level correlates of IPV among middle-aged and older adults.

## Methods

The overall aim of this analysis was to investigate IPV in middle-aged and older adults using a subset of data from the International Sexual Health and Reproductive Health (I-SHARE) conducted during COVID-19. Cross-sectional, country-level data using website data verified by academic or international health organizations were collected along with the survey.

We adapted a social-ecological theory to reflect the factors captured at the individual, relationship/household, community, and country levels (10) (S1 Table).

### Study Design (I-SHARE)

The I-SHARE survey was a cross-sectional online survey administered in 30 low-, middle-, and high-income countries between July 2020 and February 2021 (17). I-SHARE partnered with national family planning, academic, and non-profit groups in each country as well as global partners like the United Nation Family Planning Association, to support the survey implementation and establish trust in the research (16). Participants of the I-SHARE survey were at least 18 years old, a current resident of the country where they completed the survey, and able to provide online informed consent (16). Participants could stop participating or skip any questions they wished. No identifiable data was collected.^40^ Country studies linked survivors to local IPV resources.

Each in-country lead researcher organised translation to local languages, field testing, and ethical review (16). Surveys were piloted for translation on sensitive topics. The I-SHARE survey was distributed in each country through local partner organisations, sexual and productive health networks, email listservs, and social media groups determined by the in-country research lead (16). Due to varying COVID-19 restriction measures, different countries adopted sampling methods that were most contextually feasible at the time being. Twenty-three countries used convenience sampling by distributing the survey through social media, email listservs, sexual and reproductive health networks, and other non-profit and academic partners identified by in-country researchers. Six countries used online panel sampling based on key sociodemographic characteristics identified by in-country researchers and had varying degrees of population representativeness (16). Two countries used population-representative samples with frames identified by in-country researchers (S2 Table). All surveys were conducted online on personal devices.

The survey captured information on sociodemographic, relationships, sexual health and behaviour, intimate partner violence, and mental health during COVID-19 social distancing measures. Survey questions used a mix of existing validated and newly developed scales (16). To define the time period of interest for survey items which ask participants about their experiences “during social distancing measures,” I-SHARE researchers determined the start date of social distancing measures in each country based on local policies. Less than 1% of people had been socially distancing for 3 months or less at the time of completing the survey. About 20% had been socially distancing for 3 to 6 months, over 56% for 6 to 9 months, about 15% for 9 months to 12 months, and 10% for over 12 months. Each country survey underwent one to three rounds of testing (16).

### Variables selection and management

Variables were selected from each level of the social-ecological theory for analysis. Associated variables and confounders identified in the literature (S1 Table) were considered potential correlates in this study.

#### Outcome: Intimate partner violence

The I-SHARE survey measured physical, sexual, psychological, and financial IPV during COVID-19 social distancing measures using six questions. Of the six questions, five were operationalised in this study to construct psychological, sexual, and physical violence experienced during the COVID-19 social distancing measures, in congruence with the WHO validated Violence Against Women (VAW) Instrument (18) (S3 Table). Physical violence was captured from the question, ‘Has a partner slapped, pushed, hit, kicked or choked you or thrown something at you that could hurt you?’ Sexual violence was a composite of responses to two questions which captured forced sexual intercourse and sexual coercion: ‘Has a partner physically forced you to have sexual intercourse when you did not want to?’ and ‘Has a partner made you have sexual intercourse when you did not want to because you were afraid of what your partner might do?’ Psychological violence was a composite of responses to two questions which captured controlling behaviours and emotional abuse: ‘Has a partner tried to restrict (online or phone) contact with your family?’ and ‘Has a partner insulted you or made you feel bad about yourself?’ Financial violence was excluded because it was not part of the validated WHO instrument (19). Participants could choose between the responses “No,” “Yes, once,” or “Yes, multiple times” for all the violence questions.

A composite IPV variable was created for people who answered all five questions regarding physical, sexual, or psychological violence during the COVID-19 social distancing measures, from July 2020 to February 2021. If people answered ‘yes’ to any of the questions, they were coded as having experienced IPV during the COVID-19 social distancing measures.

#### Correlates at each social-ecological level

Several variables were recoded for analysis to reduce data sparsity issues. Details on the I-SHARE 2020-21 survey are available in S5 Table.

##### Individual level

Age was collected in the data as a continuous variable but was recoded into age groups, capturing people who are 45-54, 55-64, and 65 or more years old. Sex assigned at birth was collected as ‘male’, ‘female’, or ‘other’. Gender was captured as ‘cisgender,’ ‘non-cisgender,’ and ‘other.’ Those who selected ‘other’ (<1%) were regrouped with ‘non-cisgender.’ Sexual orientation was collected as a categorical variable of ‘heterosexual’, ‘bisexual’, ‘gay’, ‘lesbian’, ‘questioning or unsure’, ‘asexual’, ‘pansexual’, and ‘other’. Very few respondents identified as a sexual minority, so it was recoded as ‘heterosexual’ and ‘other sexual orientation.’ Education level was collected as a categorical variable and recoded based on the UNESCO International Standard Classification of Education (ISCED) categories.^48^ Employment status was recoded as ‘unemployed’, ‘employed’ which included those who were self-employed or informal workers, ‘retired’, and ‘other’ which included those who were students (<1%). Isolation due to COVID-19 was captured as a binary ‘yes’ or ‘no’ response. Ethnic minority status was recoded as a binary outcome by the I-SHARE team based on the in-country demographics of the participant.

##### Household and community level

Cohabitation was captured as part of the responses to a question regarding cohabitation and relationship status and recoded to a binary response to only reflect the responses regarding cohabitation. Food insecurity during COVID-19 was recoded to be a binary response: ‘Yes, more than before’ or ‘No or less than before’. At the community level, residential area was captured as a binary ‘urban’ or ‘rural’ response.

##### Country level

At the country level, social distancing stringency was created by categorising the Oxford COVID-19 Government Response Tracker (Ox-CGRT) score of each country as ‘low’ if the score was less than 50 and ‘high’ if the score was greater than 50 (20). The cut off was determined based on the median stringency score. The measure of COVID-19 social distancing stringency used in this analysis is the mean stringency that the country experienced in the duration of time from when the country’s social distancing measures began to when the surveys were administered. The start of country social distancing measures was determined by I-SHARE in-country researchers.

Gender Inequality Index (GII) scores were recoded as a binary outcome with scores less than or equal to 0.25 as high gender equality and scores from 0.25-0.5 as low gender equality. All countries had GII scores within 0-0.5. The cut off was selected based on the median. The most recent GII scores from 2019 represent the gender inequality of each country in the data.

The 2020 Social Progress Index (SPI) was chosen to represent social and environmental protection progress for each country in the study. The range of SPI scores was between about 50-100, so social progress was recoded as a binary with scores less than or equal to 75 as ‘low progress’ and scores higher than 75 as ‘high progress.’ The cut off was selected based on the median.

Country income was based on World Bank 2019-20 criteria. Low-(<$1,085) or lower-middle income ($1,086-4,255) economies were Nigeria, Lebanon, and Mozambique. Upper-middle income ($4,256-13,205) economies were Argentina, Botswana, Colombia, Mexico, Moldova, and Malaysia. High-income (>$13,205) economies were Australia, Canada, Czech Republic, Denmark, France, Germany, Italy, Latvia, Luxembourg, Panama, Portugal, Singapore, Spain, Uruguay, and the United States.

### Data inclusion and analysis

All data management and analysis were performed using STATA/SE 17.0. All individual, household, community, and country-level variables described above were tabulated. The prevalence of overall IPV and subtypes of IPV was tabulated. Country social distancing stringency, gender inequality, social progress, and country income level were tabulated to describe country level characteristics. We compared the characteristics of those missing data on any of the IPV variables to that of the whole sample and found them to be broadly similar (S6 Table A).

Of the 23067 participants in the I-SHARE survey, 4454 people were >= 45 years old. Participants who had missing data for key variables—age, sex at birth, sexual orientation, relationship status, education level, and geographic area, and any of the five questions used to construct IPV outcome (S3 Table) were excluded from this analysis.

Twenty-four countries were represented in this study—Argentina, Australia, Botswana, Canada, Colombia, Czech Republic, Denmark, France, Germany, Italy, Latvia, Lebanon, Luxembourg, Malaysia, Mexico, Moldova, Mozambique, Nigeria, Panama, Portugal, Singapore, Spain, Uruguay, and United States. The final sub-population included in the descriptive analysis (N=2867) were middle-aged and older adults who had completed the key sociodemographic items and all five IPV variables (S4 Fig).

We conducted descriptive analysis of these data to understand prevalence of IPV among this sub-population and relevant characteristics. Unadjusted odds ratios of IPV among all potential correlates were explored using logistic regression models with a random intercept for country to account for within-country clustering.

To run a fully adjusted model, people with missing data on gender, social isolation, or food insecurity were dropped by STATA due to modelling requirements. 1730 individuals from 20 countries were included for regression modelling (S2 Table). The multivariable logistic regression model included age, sex at birth, gender, sexual orientation, education level, employment status, social isolation, residential area, cohabitation status, food insecurity, country social distancing stringency level, gender inequality, social progress, and country income level were all identified as potential correlates (S4 Fig). All potential correlates were included in the final model to avoid over-fitting.^49^ The random-intercept logistic regression model was fitted with Gauss-Hermite quadrature approximation. Collinearity was checked using a backward-deletion strategy by removing correlates individually and assessing whether the confidence intervals of the remaining correlates narrowed in the model (21).

Correlates were only removed from the model if there was evidence of collinearity. No collinearity was found so all correlates were included in the final model. Likelihood ratio tests were performed to obtain a global p-value for each correlate.

Two sensitivity analyses were conducted, first to test if the model produced similar results in middle-aged and older adults in low- and middle-income countries (LMIC), and second with IPV defined by only physical and sexual violence (17).

### Ethical Approval

The I-SHARE survey received local ethical approval and institutional review board approval in each country. A data sharing agreement was signed by all collaborating institutions.

Participants provided informed consent and no identifying data was collected. This study was approved by the London School of Hygiene & Tropical Medicine Ethics Committee.

## Results

### Sample characteristics

Data from 2867 middle-aged and older adults in 24 countries were analysed. The majority of participants were college or university educated (63.7%), heterosexual (86.1%), cisgender (88.6%), females (55.4%), and between the ages of 45-54 (53.7%). Three-quarters were employed and two-thirds lived in an urban area. During COVID-19, most people lived with a partner (77.2%), did not isolate due to COVID-19 (84.7%), and did not experience increased food insecurity (82.9%) (Table 1).

**Table 1:** Description of study sample characteristics from I-SHARE 2020-21 (N=2867 unless otherwise specified).

|  |  | **N\*** | **%**<br>**(Column %)** | **% of IPV**<br>**(Row %)** |
| --- | --- | --- | --- | --- |
| **Age group in years** | 45-54 | 1539 | 53.7 | 14.6 |
|  | 55-64 | 824 | 28.7 | 11.5 |
|  | ≥ 65 | 504 | 17.6 | 5.6 |
| **Sex at birth** | Male | 1278 | 44.6 | 11.3 |
|  | Female | 1589 | 55.4 | 12.8 |
|  | Other<sup>1</sup> | 0.0 | 0.0 | 0.0 |
| **Gender**<br>**(N=2857)** | Cisgender | 2531 | 88.6 | 11.7 |
|  | Non-cisgender | 326 | 11.4 | 15.6 |
| **Sexual orientation** | Heterosexual | 2469 | 86.1 | 12.0 |
|  | Other sexual orientation<sup>2</sup> | 398 | 13.9 | 13.1 |
| **Ethnic minority status<sup>3</sup>**<br>**(N=1509)** | Minority | 197 | 13.1 | 24.4 |
|  | Not ethnic minority | 1312 | 86.9 | 10.1 |
| **Education level** | No formal and primary | 70 | 2.4 | 10.0 |
|  | Secondary | 755 | 26.3 | 8.0 |
|  | College/University | 1826 | 63.7 | 13.5 |
|  | Other<sup>1</sup> | 216 | 7.5 | 16.2 |
| **Employment status** | Employed | 2154 | 75.1 | 12.9 |
|  | Unemployed | 83 | 2.9 | 14.5 |
|  | Retired | 526 | 18.4 | 6.1 |
|  | Other<sup>1</sup> | 104 | 3.6 | 25.0 |
| **Residential area** | Rural | 944 | 32.9 | 8.7 |
|  | Urban | 1923 | 67.1 | 13.8 |
| **Cohabitation status** | Not living with partner | 653 | 22.8 | 14.6 |
|  | Living with partner | 2214 | 77.2 | 11.4 |
| **Ever isolated due to**<br>**COVID-19 (N=2859)** | No | 2422 | 84.7 | 11.4 |
|  | Yes | 437 | 15.3 | 16.5 |
| **Food insecurity during**<br>**the pandemic (N=1741)** | No or less than before | 1444 | 82.9 | 13.5 |
|  | Yes more than before | 297 | 17.1 | 26.9 |
| **Country social**<br>**distancing stringency** | Low stringency | 897 | 31.3 | 12.7 |
|  | High stringency | 1970 | 68.7 | 11.9 |
| **Gender inequity index (GII)<sup>5</sup>** | Low equality | 913 | 31.9 | 18.8 |
|  | High equality | 1954 | 68.2 | 9.0 |
| **Social progress index (SPI)<sup>6</sup>** | Low progress | 570 | 19.9 | 22.1 |
|  | High progress | 2297 | 80.1 | 9.7 |
| **World Bank country income level<sup>7</sup>** | High | 2118 | 73.9 | 9.4 |
|  | Upper-middle | 735 | 25.6 | 19.7 |
|  | Low or lower-middle | 14 | 0.5 | 21.4 |

\* Numbers may differ from the total sample size of 2867 due to missing values.
<sup>1</sup> "Other" was a survey response option. Participants were unable to specify further.
<sup>2</sup> "Other sexual orientation" includes people who responded as bisexual, gay, lesbian, questioning or unsure, asexual, pansexual, and other.
<sup>3</sup> Individual's ethnic minority status was identified based on their country demographics.
<sup>4</sup> Country social distancing stringency levels were based on the Ox-CGRT score. Countries with $\leq 50$ had low stringency and countries with $>50$ had high stringency.
<sup>5</sup> GII measures gender inequalities within a society. Scores $\leq 0.25$ had the most gender equality and scores from 0.25-0.5 had low equality.
<sup>6</sup> SPI measures the social and environmental progress of a country. Scores $\leq 75$ were considered low and $>75$ was high.
<sup>7</sup> Countries were grouped based on World Bank 2019-20 criteria.

Half of the participants resided in a country with a high social distancing policy stringency level (68.7%) and the majority were from countries with higher gender equality (68.2%) and greater social progress (80.1%). More participants were in high-income countries compared to low and middle-income countries (Fig 1).

**Fig 1:**
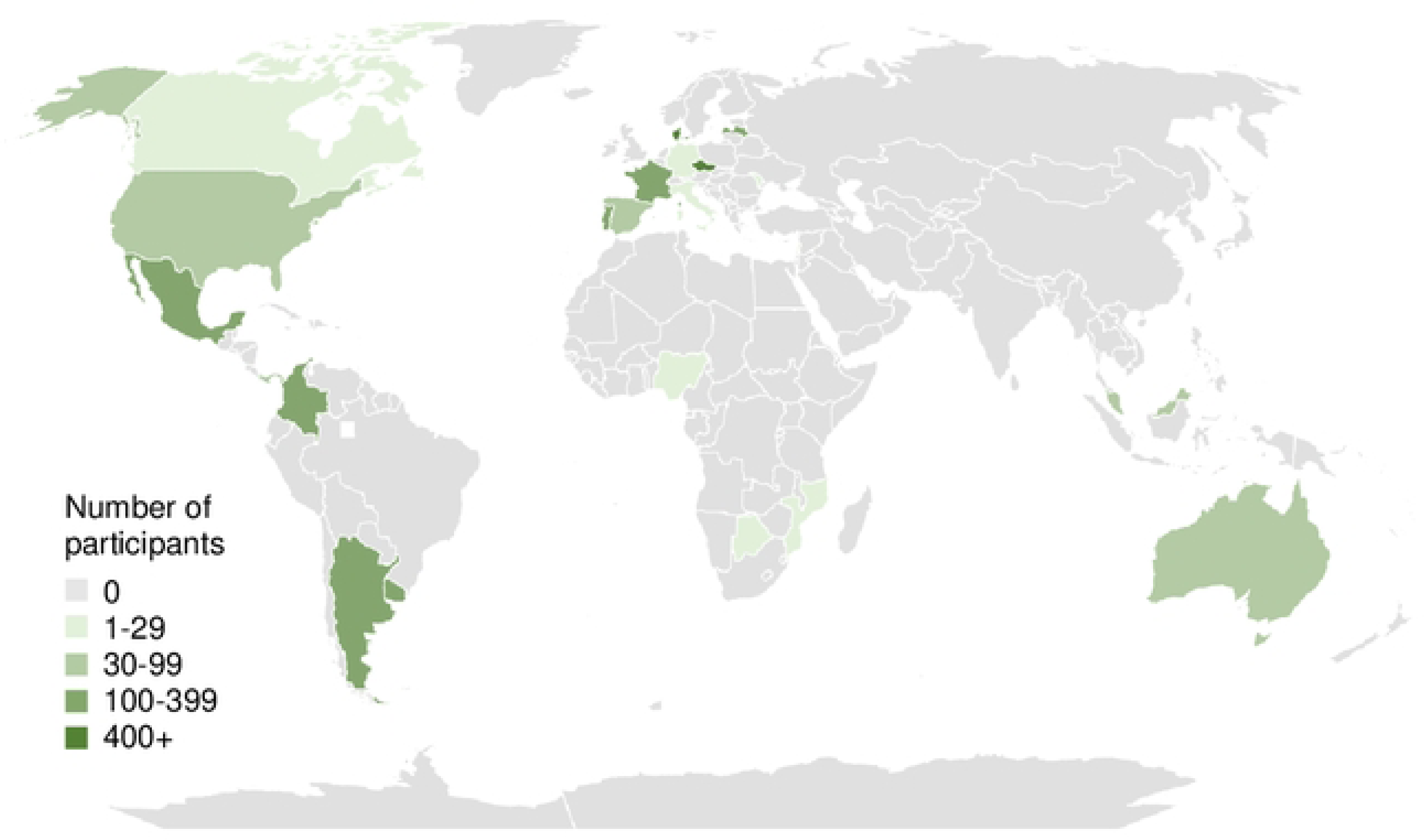
Countries with middle-aged and older adults from the I-SHARE 2020-21 survey included in this study (N=2867).

### Prevalence of overall and subtypes of IPV

Among the 2867 participants in the study 12.1% (346/2867) of people experienced some form of IPV during COVID-19 social distancing measures (Table 2). Ethnic minorities (368/1509) reported more than twice the prevalence of IPV compared to people who were not ethnic minorities (152/1509). People who experienced more food insecurity during the pandemic experienced almost twice the amount of IPV (468/1741) compared to those who did not (235/1741). IPV was also lower in countries with high gender equality (258/2867), social progress (278/2867), and income (269/2867). The frequency of IPV appeared to be almost equal between countries with high (341/2867) and low (364/2867) social distancing stringency levels (Table 1).

**Table 2:**
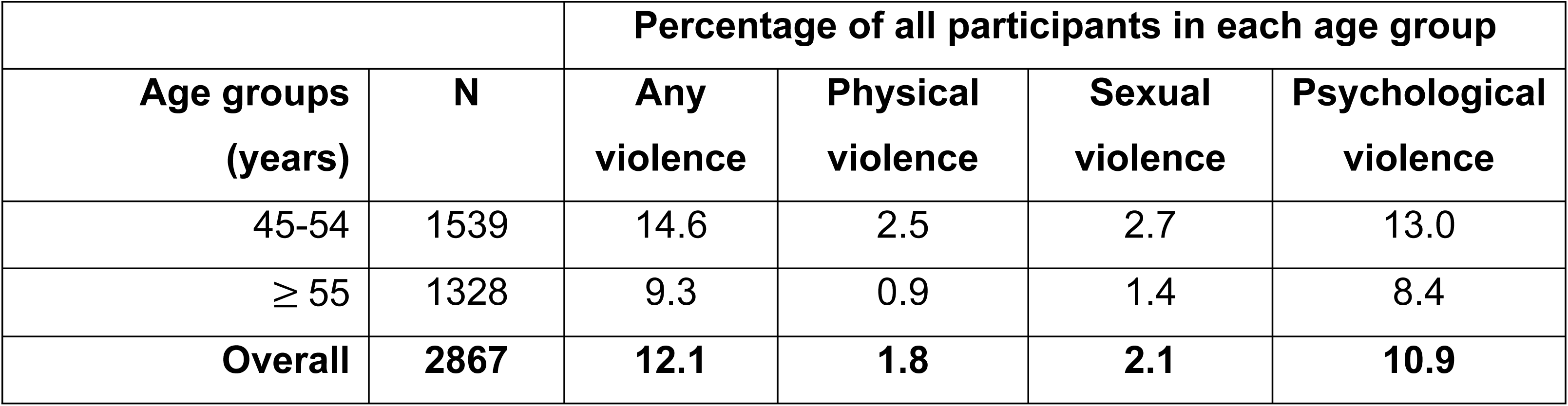
The prevalence of overall IPV and subtypes of IPV, including physical, sexual, psychological violence across the age groups from the I-SHARE 2020-21 survey (N=2867).

|  |  | Percentage of all participants in each age group |  |  |  |
| --- | --- | --- | --- | --- | --- |
| Age groups (years) | N | Any violence | Physical violence | Sexual violence | Psychological violence |
| 45-54 | 1539 | 14.6 | 2.5 | 2.7 | 13.0 |
| $\geq 55$ | 1328 | 9.3 | 0.9 | 1.4 | 8.4 |
| **Overall** | **2867** | **12.1** | **1.8** | **2.1** | **10.9** |

The prevalence of overall and subtypes of IPV was lower in people aged 55 or older compared to those aged 45-54 years (Table 2). Psychological violence was experienced by 10.9% (312/2867) of participants. It was more prevalent than physical violence (1.8%, 52/2867) and sexual violence (2.1%, 60/2867). Participants in the oldest group (≥ 55) reported 6 times more psychological violence was 6 times than sexual violence and 9 times more than physical violence. Among people 45-54 years, psychological violence was experienced 5 times more than physical and sexual violence. (Table 2).

Among the 348 people who reported experiencing IPV during the COVID-19 pandemic, 16.4% (57/348) experienced two or more forms of violence and 5.5% (19/348) experienced all three subtypes of IPV (Fig. 2). 14.7% (51/348) experienced physical violence and 2.9% (10/348) experienced physical violence as their only form of IPV. 17.5% (61/348) reported experiencing sexual violence and 7.1% (25/348) experienced only sexual violence. 89.6% (312/348) experienced psychological violence by their partners and 73.5% (256/348) experienced only psychological violence (Fig. 2). Psychological violence was not only the most prevalent subtype of IPV, but also the most frequently co-occurring subtype with the others among middle-aged and older adults who experienced violence during the pandemic.

**Fig 2:**
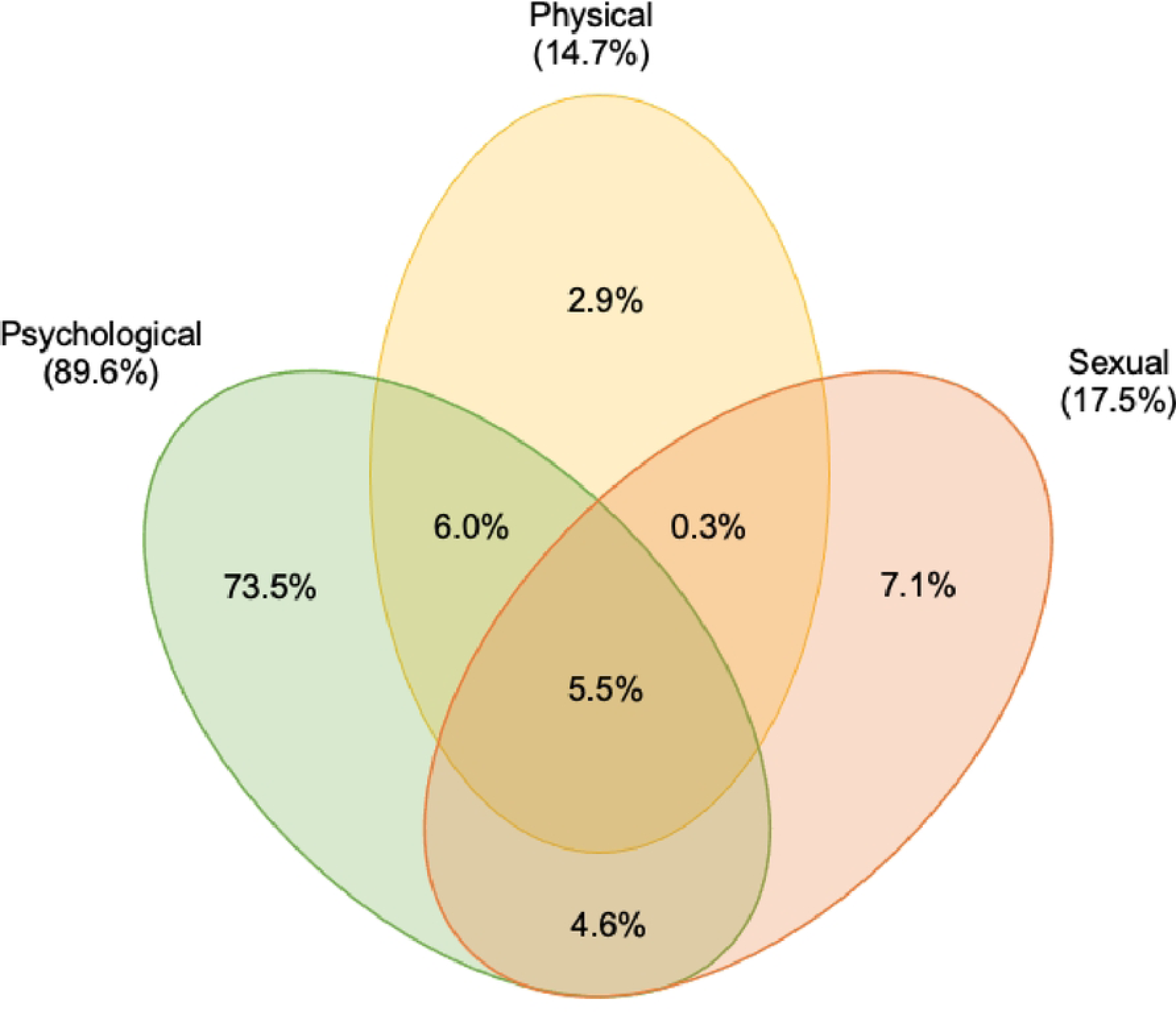
Proportion of each subtype and overlapping subtypes of IPV experienced among middle-aged and older adults during the pandemic in the I-SHARE 2020-21 survey (N=348). Areas are not proportional to the values. Adapted from Yoshihama and Sorenson (1994).^52^

Of the 16.4% (57/348) who experienced multiple forms of violence, 98.2% of them (56/57) experienced psychological violence. 10.4% (36/348) experienced sexual violence and 11.8% (41/348) experienced physical violence.

Physical and sexual violence are often conceptualised as commonly co-occurring with one another. Of those experiencing physical violence, 40% also experienced sexual violence (20/51). Of those experiencing sexual violence, a third also experienced physical violence (20/61).

### Identifying individual, household, community, and country level correlates

Age group, sex at birth, sexual orientation, and education level did not appear to be associated with IPV in bivariate analyses accounting for clustering by country. At the individual level, people who ever isolated due to COVID-19 were more likely to have experienced IPV compared to those who did not after accounting for clustering by country (clustered OR (cOR)=1.5, 95% CI 1.1, 2.0). At the household level, those who experienced more food insecurity were more likely to have experienced IPV compared to those who did not have increased food insecurity (cOR=2.4, 95% CI 1.8, 3.3). At the country level, people who lived in high social distancing stringency countries (cOR=0.5, 95% CI 0.3, 0.8) or highly socially progressive countries (cOR=0.6, 95% CI 0.4, 0.9) were less likely to have experienced IPV compared to their counterparts.

The final model, which adjusted for all other correlates and accounted for clustering by country, showed that some of the above associations remained significant. Ever being isolated (aOR=1.4, 95% CI 1.0, 2.0), increased food insecurity during the pandemic (aOR=2.2, 95% CI 1.6, 3.0), and living in countries with a high level of social distancing stringency (aOR=0.6, 95% CI 0.4, 0.9) remained significant associated factors. But social progressiveness, gender inequality and country income level were not found to be associated with IPV in the final model (Table 3).

**Table 3:** Unadjusted and adjusted association between correlates and IPV during COVID-19 social distancing among middle-aged and older adults in the I-SHARE 2020-21 survey (N=1730)^1^.

|  |  |  | Unadjusted OR<br>(95% CI)<sup>2</sup> | Final model:<br>aOR<sup>3</sup> (95% CI) | Global P-value<sup>4</sup> |
| --- | --- | --- | --- | --- | --- |
| Individual Level | Age group in years | 45-54 | 1 | 1 | 0.54 |
|  |  | 55-64 | 0.8 (0.6, 1.1) | 0.9 (0.6, 1.2) |  |
|  |  | ≥ 65 | 0.7 (0.4, 1.1) | 0.7 (0.4, 1.4) |  |
|  | Sex at birth | Male | 1 | 1 | 0.40 |
|  |  | Female | 1.0 (0.8, 1.3) | 0.9 (0.7, 1.2) |  |
|  | Gender | Cisgender | 1 | 1 | 0.18 |
|  |  | Non-cisgender | 1.2 (0.8, 1.9) | 1.3 (0.9, 2.0) |  |
|  | Sexual orientation | Heterosexual | 1 | 1 | 0.97 |
|  |  | Other sexual orientation | 1.0 (0.7, 1.4) | 1.0 (0.7, 1.5) |  |
|  | Education level | No formal and primary | 1 | 1 | 0.89 |
|  |  | Secondary | 0.8 (0.3, 2.2) | 1.0 (0.4, 2.6) |  |
|  |  | College/University | 0.9 (0.3, 2.3) | 1.1 (0.4, 2.5) |  |
|  |  | Other | 1.0 (0.3, 2.9) | 1.2 (0.4, 3.3) |  |
|  | Employment status | Employed | 1 | 1 | 0.03 |
|  |  | Unemployed | 1.3 (0.7, 2.5) | 1.2 (0.6, 2.4) |  |
|  |  | Retired | 0.8 (0.5, 1.4) | 1.1 (0.6, 2.0) |  |
|  |  | Other | 2.3 (1.4, 3.8) | 2.2 (1.3, 3.7) |  |
|  | Ever isolated due to COVID-19 | No | 1 | 1 | 0.04 |
|  |  | Yes | 1.5 (1.1, 2.0) | 1.4 (1.0, 2.0) |  |
| Household & Community Level | Cohabitation status | Not living with partner | 1 | 1 | 0.81 |
|  |  | Living with partner | 1.0 (0.7, 1.4) | 1.1 (0.8, 1.4) |  |
|  | Food insecurity during the pandemic | No or less than before | 1 | 1 | <0.0001 |
|  |  | Yes more than before | 2.4 (1.8, 3.3) | 2.2 (1.6, 3.0) |  |
|  | Residential area | Rural | 1 | 1 | 0.80 |
|  |  | Urban | 1.2 (0.8, 1.7) | 1.0 (0.7, 1.5) |  |
| Country Level | Country social distancing stringency | Low stringency | 1 | 1 | 0.04 |
|  |  | High stringency | 0.5 (0.3, 0.8) | 0.6 (0.4, 0.9) |  |
|  | Gender inequality index (GII) | Low equality | 1 | 1 | 0.41 |
|  |  | High equality | 0.7 (0.5, 1.1) | 0.8 (0.5, 1.3) |  |
|  | Social progress index (SPI) | Low progress | 1 | 1 | 0.14 |
|  |  | High progress | 0.6 (0.4, 0.9) | 0.4 (0.1, 1.4) |  |
|  | World Bank country income level | High | 1 | 1 | 0.49 |
|  |  | Upper-middle | 1.4 (0.8, 2.3) | 0.5 (0.2, 1.7) |  |
|  |  | Low or Lower-middle | 1.7 (0.4, 6.7) | 0.7 (0.1, 4.2) |  |

<sup>1</sup> Total N is lower due to missing data.
<sup>2</sup> Accounts for clustering by country.
<sup>3</sup> Adjusted for all other covariates. Accounts for clustering by country. The final model includes data from 20 countries—Australia, Canada, Colombia, France Germany, Italy, Latvia, Lebanon, Luxembourg, Malaysia, Mexico, Moldova, Mozambique, Nigeria, Panama, Portugal, Singapore, Spain, Uruguay, and USA.
<sup>3</sup> This is the p-value for the final model. Global p-values were determined by likelihood ratio tests.
Note: Ethnic minority status was excluded in regression models due to missing values.

### Sensitivity Analyses

A sensitivity analysis among the 749 participants from LMIC resulted in the same overall associations between social isolation (aOR 1.8, 95%CI 0.6, 1.7, p=0.02), food insecurity (aOR 3.4, 95% CI 1.5, 3.8, p=0.0004), and country social distancing stringency level (aOR 0.5, 95% CI 0.3, 0.8, p=0.02) (S7 Table A). A sensitivity analysis among 2867 participants to operationalise IPV with another standard definition which only includes physical and sexual violence resulted in similar directions and effect sizes for social isolation (aOR 1.6, 95% CI 0.9, 2.8, p=0.13), food insecurity (aOR 2.0, 95% CI 1.2, 3.5, p=0.02), and country social distancing stringency level (aOR 0.5, 95% CI 0.3, 1.0, p=0.06) compared to the final model (S7 Table B). However, fewer correlates appeared to be associated with this IPV outcome.

## Discussion

12.1% of middle-aged and older adults reported experiencing IPV in the period between their local social distancing measures and the I-SHARE survey. The prevalence of IPV was the same between males and females and decreased as age group increased. Psychological violence was the most common subtype of violence. Increased isolation, increased food insecurity and lower social distancing stringency were associated with greater IPV among middle-aged and older adults. This secondary analysis of I-SHARE 2020-21 extends the literature by focusing on middle-aged and older adults using multi-country data.

The overall prevalence of IPV during COVID-19 among middle-aged and older adults estimated by this study was found to be higher than the 7.0% (1,070/15,336) of participants from the whole I-SHARE sample who reported experiencing IPV. The most common form of IPV across all middle and older age groups was psychological violence. This is consistent with the existing literature on IPV during COVID-19 (22–26) and broader patterns seen in IPV (27–29). Psychological violence was found to overlap with physical and sexual subtypes of IPV. Previous studies suggest that psychological violence is an important risk factor for other types of violence (27,28).

Several studies have highlighted the extreme toll that psychological violence has on mental distress (30,31), self-esteem, feeling worthless, loss of identity, as well as a higher prevalence of post-traumatic stress disorder (PTSD) and complex-PTSD (29,32–34).

Unfortunately, middle-aged and older people experiencing IPV during the pandemic also had reduced access to IPV support services (35) and general mental health resources (36). Low digital connectivity among this age group has also made many remote service replacements inaccessible, leaving them without access to necessary support (37).

There was strong evidence that having isolated due to COVID-19 was associated with increased odds of IPV compared to those who did not isolate among middle-aged and older adults. This finding coincides with a broader base of literature that identifies social isolation as a risk factor for IPV (38–40) and conversely that social support is protective (41). During the pandemic, many people self-isolated (42). There are several possible explanations for the observed association between social isolation and experiences of IPV. Isolation leads to weakened social networks, decreasing help-seeking opportunities while increasing a perpetrator’s ability to control and coerce survivors (38). Additionally, perpetrators could blame survivors who are isolating due to symptoms or exposure to COVID-19 for putting them at risk of infection, triggering heightened stress, anxiety, and poor mental health in both survivors and perpetrators, which could lead to new or escalating forms of IPV (22,26,43).

Higher odds of IPV were observed in people who experienced increased food insecurity during the COVID-19 pandemic compared to those who did not. The association between food insecurity and IPV has been well established in many countries (44–46). Food insecurity is often discussed in the context of a general lack of material security, such as housing or economic insecurity (44–46). Stress theory suggests that insufficient or limited resources at the individual and household level is a source of tension that can increase risk of IPV (45–47). It posits that stress associated with too much demand over too little resources leads to IPV (47).

We found lower odds of IPV in countries with higher social distancing stringency. Current literature on the effect of social distancing policies on IPV has produced mixed evidence on its relationship with IPV (13), and in many previous studies, social distancing was found to be associated with an increased prevalence of IPV while simultaneously keeping IPV hidden (5,48,49). Our study findings among middle-aged and older adults were inconsistent with most of these previous findings. One potential explanation may be that these countries with more stringent social distancing policies might also be more likely to implement stronger social protections (50). Literature on social protection during COVID-19 shows that cash transfers were the most commonly implemented social protections, as well as food vouchers and wage subsidies (51). These protections were primarily implemented by high-income countries alongside social distancing policies (51). Without indexes that capture the impact of concurrent COVID-19 policy responses, the relationship between social distancing stringency, social protections, and IPV cannot be disaggregated.

Our study has several limitations. We used a cross-sectional online survey, which is unable to determine the direction of association between correlates and outcomes. As a result, causal claims cannot be made for any of the correlates identified in this study. Participants may not be comfortable disclosing experiences of IPV which could lead to an underestimation of the prevalence of IPV.

Missing data from this study could impact the generalisability of the findings and is a limitation. About 30% of people had missing IPV outcome data. However, people with missing data for IPV had similar sociodemographic characteristics to those people who were included in the descriptive analysis. Efforts were made to increase the generalisability of these findings by using diverse sampling methods in the I-SHARE survey, such as including population representative sampling frames and using online panels (S2 Table). Sensitivity analysis showed that the model is also sensitive across LMIC subpopulations, with the same directions of associations and larger effect sizes compared to the final model (S7 Table A).

Further studies are needed to understand the relationship between country level variables, like social distancing stringency, and the pathways through which they impact IPV in various country settings. Future research to evaluate the impacts of social distancing and social protection policies on IPV during COVID-19 are needed. Until now, research on violence during COVID-19 considers single policy measures. Intimate partner violence and the intersection of aging is a complex phenomenon that needs to be understood in the context of suites of pandemic and crisis policies, which interact in multifaceted ways to shape experiences of violence. Detailed examination of mediation effects and cross-level interactions within the social-ecological theory is needed to continue to expand the understanding of how correlates are associated with IPV.

## Declarations

### Ethics approval and consent to participate

The I-SHARE survey received local ethical approval and institutional review board approval in each country. A data sharing agreement was signed by all collaborating institutions.

Participants provided informed consent and no identifying data was collected. This study was approved by the London School of Hygiene & Tropical Medicine Ethics Committee.

### Consent for publication

Not applicable.

### Availability of data and materials

The survey protocol and final survey instrument are available. Requests for data access must follow the rules outlined in the I-SHARE data sharing agreement. Data code can be made available upon request to the corresponding author.

### Authors’ contributions

GC, JT, DW, and CC conceptualized the study. GC wrote the first draft of the manuscript. GC completed the data analysis with support from JT, KW, and DW. All authors critically revised the article for important intellectual content and approved the final version. All authors contributed to the knowledge generated, article processing fees, and are available for future correspondences regarding the paper. All authors had full access to all of the data and had final responsibility for the decision to submit for publication.

## Data Availability

Data for this study are available upon request. The full I-SHARE dataset is still being analyzed by the team and not yet ready for open access. Requests can be considered by the I-SHARE research consortium if a proposal is submitted to the I-SHARE team for assessment.

## Acknowledgements

We want to acknowledge the following I-SHARE members for their contributions to this article: Kristien Michielsen, Vinicius Jobim Fischer, Amr Abdelhamed, Noor Ani Ahmad, Juliana Anderson, Nicholás Brunet, Sharyn Burns, Leonardo Chavane, Juan Carlos Rivillas, Fiorella Farje De la Torre, José de Jesús González-Salazar, Gert Martin Hald, Devon Hensel, Corina Iliadi-Tulbure, Olena Ivanova, Anna Kågesten, Kateřina Klapilová, Lucia Knight, Dorie Kogut, Eneyi Kpokiri, Gunta Lazdane, Alejandra Lopez-Gomez, Ismael Maatouk, Filippo Maria Nimbi, Caroline Moreau, Rocio Murad Rivera, Viola Nilah Nyakato, Pedro Nobre, Caitlin Alsandria O’Hara, Shania Pande, Emilie Peeters, Carles Pericas, Lore Remmerie, Juan Rivillas, Eusebio Rubio-Aurioles, Osama Shaeer, Jenna Marie Strizzi, Kun Tang, Inês M. Tavares, Jennifer Toller Erausquin, Sonam Shah, Eline Van Damme, Wah Yun Low, and WeiHong Zhang. A full list of I-SHARE consortium members can be found on the I-SHARE website. We would like to thank the Academic Network for Sexual and Reproductive Health and Rights Policy, Social Entrepreneurship to Spur Health, the London School of Hygiene and Tropical Medicine Sexually Transmitted Research Interest Group, and the University of North Carolina at Chapel Hill Institute for Global Health and Infectious Diseases.

## Supporting Information

**S1 Table: Potential correlates identified from literature on IPV during COVID-19 organised into the four-level social-ecological theory adapted from Dahlberg and Krug** (**2006**). Items in bold and italics were collected in I-SHARE 2020-21 or publicly available through country-level indexes. Items in bold were included in the final model (except for ethnic minority status). Items in italics were conceptualised as mediators and therefore not included in the final model.

**S2 Table: Number and percentage of countries in I-SHARE 2020-21 by sampling strategy.** Low-(<$1,085) or lower-middle income ($1,086-4,255) economies were Nigeria, Lebanon, and Mozambique. Upper-middle income ($4,256-13,205) economies were Argentina, Botswana, Colombia, Mexico, Moldova, and Malaysia. High-income (>$13,205) economies were Australia, Canada, Czech Republic, Denmark, France, Germany, Italy, Latvia, Luxembourg, Panama, Portugal, Singapore, Spain, Uruguay, and the United States.

**S3 Table: I-SHARE 2020-21 adaptations of the WHO Violence Against Women (VAW) questions used to construct IPV.**

**S4 Fig: Descriptive analysis population (N=2867) and bivariate and final model population (N=1730) selection parameters from I-SHARE 2020-21.**

**S5 Table: Questions on the I-SHARE 2020-21 survey used for the analysis.**

**S6 Table A: Characteristics of people missing and not missing IPV outcome responses in the study from I-SHARE 2020-21 (N=4057).**

**S6 Table B: Characteristics of people excluded from the final model due to missing data on gender, isolation, and food insecurity from I-SHARE 2020-21 (N=2867).**

**S7 Table A: A sensitivity analysis with participants from LMICs to determine if the model is sensitive within this population from the I-SHARE 2020-21 survey (N=749).**

**S7 Table B: Sensitivity analysis of the construction of IPV. It is defined in this analysis as physical and sexual violence using data from the I-SHARE 2020-21 survey (N=2867).**

**S8 Table: Percentage of people in each country social distancing stringency level at each country-level variable in I-SHARE 2020-21 (N=2867).**

